# Bridging Structural MRI with Cognitive Function for Individual Level Classification of Early Psychosis via Deep Learning

**DOI:** 10.1101/2022.05.02.22274572

**Authors:** Yang Wen, Chuan Zhou, Leiting Chen, Yu Deng, Martine Cleusix, Raoul Jenni, Philippe Conus, Kim Q. Do, Lijing Xin

## Abstract

Recent efforts have been made to apply machine learning and deep learning approaches to the automated classification of schizophrenia using structural magnetic resonance imaging (sMRI) at the individual level. However, these approaches are less accurate on early psychosis (EP) since there are mild structural brain changes at early stage. As cognitive impairments is one main feature in psychosis, in this study we apply a multi-task deep learning framework using sMRI with inclusion of cognitive assessment to facilitate the classification EP patients from healthy individuals. Unlike previous studies, we used sMRI as the direct input to perform EP classifications and cognitive estimations. The proposed model does not require time-consuming volumetric or surface based analysis and can provide additionally cognition predictions. Extensive experiments were conducted on a sMRI data set with a total of 77 subjects (38 EP patients and 39 healthy controls), and we achieved 74.9±4.3% five-fold cross-validated accuracy and an area under the curve of 71.1±4.1% on EP classification with the inclusion of cognitive estimations. We reveal the feasibility of automated cognitive estimation using sMRI by deep learning models, and also demonstrate the implicit adoption of cognitive measures as additional information to facilitate EP classifications from healthy controls.

## 1. Introduction

Artificial intelligence (AI) approaches, particularly machine learning (ML) and deep learning (DL), have been extensively studied to accelerate medical data analysis and assist clinical interventions in many pathological contexts [51, 49]. Many applications have been conducted in psychiatric disorders using neuroimaging measures (*e*.*g*., sMRI [35]) as input and incorporated with AI models (*e*.*g*., supported vector machine and artificial neural networks) to establish automated diagnostic workflows at a single subject level [36, 28]. Previous machine learning works in schizophrenia have used handcrafted features extracted from sMRI data to distinguish patients from healthy individuals [12], but such feature extraction process usually involves a long computational time. To reduce computational cost, recent efforts have focused on using directly sMRI images as input, and promising results have been achieved with the help of the latest AI models (e.g., convolutional neural networks, CNNs) [38, 22]. However, these studies have mainly focused on patients at chronic stage, the classification of early psychosis (EP) patients from healthy controls (HCs), is considered to be more challenging [13, 57, 10, 48], because the brain structural changes in EP patients are mild and not evident, making computer-aided classification methods less robust and accurate.

Furthermore, progressive cognitive deficit is one major feature of schizophrenia [33, 52, 6], inspiring the possibility of using individual cognition levels, in addition to sMRI images, to facilitate automated classification of EP patients from HCs. Several recent studies have used the DL framework to incorporate cognitive estimation into the workflow to facilitate the diagnosis of Alzheimers disease by explicitly including cognitive measures as secondary inputs [53, 34]. However, this approach requires additional cognitive assessment that is not part of routine neuropsychiatric clinical examinations. Moreover, although several studies have been done using sMRI images to identify individual cognitive impairments via DL [24, 54], to the best of our knowledge, no study has been done to incorporate cognition estimation for classifying EP patients and controls.

Therefore, in this study, we applied a multi-task DL model by using sMRI as an input to classify EP patients from healthy controls and to simultaneously predict cognition levels at the single subject level. We further investigated whether the inclusion of cognitive levels estimation could facilitate the classification for EP patients and controls. Specifically, as shown in Figure 1B, a three-dimensional convolutional neural network (3D-CNN) is used to learn discriminative structural features directly from sMRI arrays. Then, three multilayer perceptron (MLP) subbranches are used to perform EP/HC classifications and cognition estimations. We have evaluated the proposed model on an in-house data set, consisting of 77 sMRI 3D arrays (38 EP patients, 39 HCs). These sMRI arrays were used to train the model in an end-to-end manner. While most of ML-based classifiers relied on features of time-consuming volumetric or surface based analysis, the proposed method performs EP/HC classifications and cognition estimation using only sMRI as input. Our major contributions can be summarized as follows.

**Figure 1:**
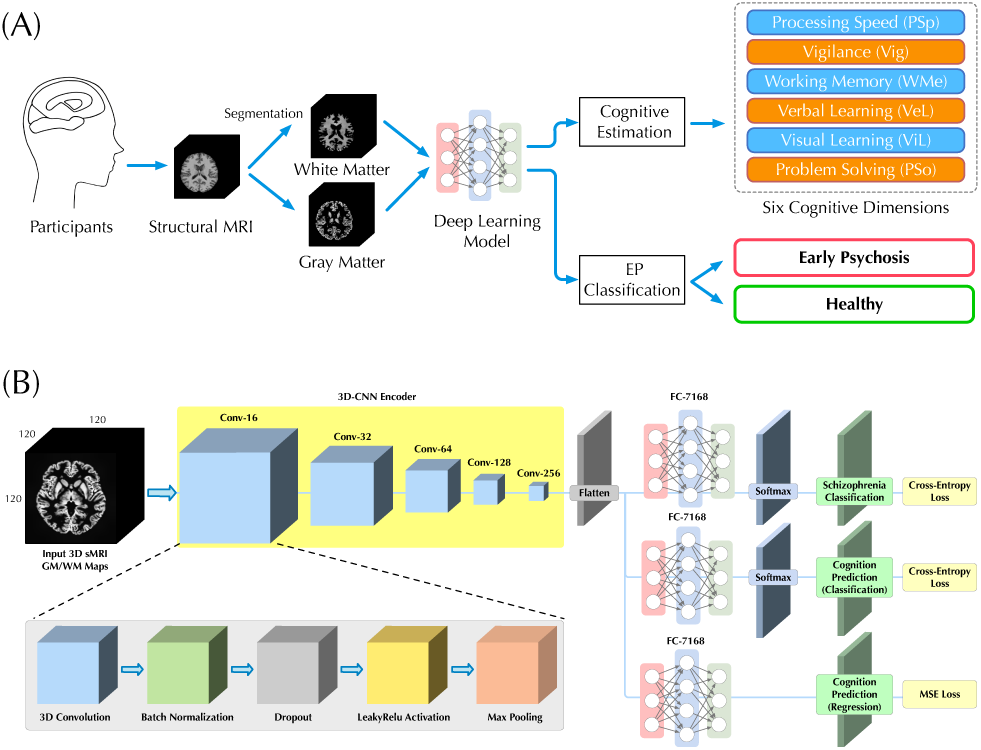
Illustrations of (A) our workflow for classification of EP patients from HCs and cognitive estimation on six dimensions, and (B) the deep learning architecture with a 3D-CNN feature encoder and three independent MLP subbranches for different subtasks of EP classification and cognitive estimations.

- A multi-task deep learning model is applied to bridge sMRI and cognitive estimation for improving the classification performance of EP, which can automatically capture structural features from 3D sMRI scans for EP classification and provide cognition as supporting evidence at individual level within a unified framework.
- The feasibility of using sMRI scans to estimate an individual’s cognitive level through deep learning was investigated.
- Extensive experimental results on in-house data set demonstrated that taking cognition measures implicitly improve classification accuracy for EP.
- The main structural contributors involved in the process of EP classification and cognitive estimation are identified.

## 2. Materials and Methods

### 2.1. Problem Setup

As shown in Figure 1A, given the sMRI image, we seek to estimate participant’s cognitive level and classify EP patients from healthy individuals in a fully automated manner. Unlike previous studies [11, 29, 26, 53], we directly utilized sMRI images as input without additional imaging analysis (*e*.*g*., voxel-based morphometry), which allowed us to more natively understand how brain structure itself contributes to the EP classification and cognition estimation.

### 2.2. Materials and Data Set

#### 2.2.1. Participants

sMRI data and corresponding neurocognitive scores were obtained from Department of Psychiatry at the Lausanne University Hospital (CHUV). The data set consists of 38 EP patients and 39 healthy controls (HC). Detailed demographic information of all participants are shown in Table 1. Specifically, the Positive and Negative Syndrome Scale (PANSS) was provided as the sum of positive, negative and general PANSS values. The patients with EP were recruited from the TIPP Program (Treatment and Early Intervention in Psychosis Program, University Hospital, Lausanne, Switzerland) [4].

**Table 1.** Demographic information of 77 subjects. PANSS, Positive and Negative Syndrome Scale.

|  | HC | EP | Total |
| --- | --- | --- | --- |
| Number of Patients | 39 | 38 | 77 |
| Gender (Female/Male) | 8/31 | 7/31 | 15/62 |
| Ethnicity (Caucasian/Other) | 28/11 | 28/10 | 56/21 |
| Age (year) | 24.54 ± 5.35 | 24.29 ± 4.23 | 24.42 ± 4.81 |
| Duration of Psychosis (year) | - | 1.00 ± 0.73 | - |
| PANSS (total) | - | 56.45 ± 13.75 | - |

All the participants provided informed written consent for this study, and the procedure was approved by the local Ethics Committee (Commission cantonale déthique de la recherché sur lêtre humain - CER-VD), in accordance with the Declaration of Helsinki. Detailed recruitment criteria for participants can be found in *Appendix: A*.

#### 2.2.2. Structural MRI Acquisition

Patients and controls underwent magnetic resonance imaging at a 7 Tesla/68 cm MR scanner (Siemens Medical Solutions, Erlangen, Germany). A 32-channel receive coil (NOVA Medical Inc., MA) with a single channel volume transmit coil was used. 3D T1-weighted MR images were acquired using MP2RAGE (TE/TR = 1.87/5500 ms, TI1/TI2 = 750/2350 ms, *α*1/*α*2 = 4°/5°, slice thickness = 1mm, FOV= 240 × 256 × 160 mm^3^, voxel size = 1mm^3^ isotropic, bandwidth = 240 Hz/Px) [32]. The original dimension of acquired sMRI data array is 240 × 256 × 160.

#### 2.2.3. Preprocessing

To generate appropriate inputs, we performed preprocessing of sMRI data using CAT12 toolkit for estimation of the probability maps of white matter (WM) and gray matter (GM). Skull striping and registration to standard space with MNI152 template were performed. Then, probability maps of WM and GM were generated after tissue segmentation and bias correction. The resulting WM and GM probability maps were down-sampled to 120 × 120 × 120 for computational efficiency. The same implementation that using only WM and GM without CSF as inputs to the model was followed as in the previous studies [].

#### 2.2.4. Neurocognitive Measures

The MATRICS Consensus Cognitive Battery (MCCB) [25, 37] was assessed for both EP and HC groups, excluding the Mayer-Salovey-Caruso Emotional Intelligence Test (MSCEIT), which does not translate well into French as an index of social cognition. The neurocognitive measures include six dimensions, *i*.*e*., processing speed (PSp), vigilance (Vig), working memory (WMe), verbal learning (VeL), visual learning (ViL) and problem solving (PSo). There exists some missing entries in the cognitive assessment data, so we replaced all missing data with the average values to generate proper training data [34]. The quantity of missing entries is: PSp 5, Vig 3, WMe 4, VeL 1, ViL 1, and PSo 1. There is at most one missing cognitive dimension per subject. The distribution of scores for all cognitive dimensions are shown in Table 2. Two-tailed student t-test was performed between the two groups, and significant difference was found on PSp, VeL and ViL with a *p* value< 0.05.

**Table 2.**
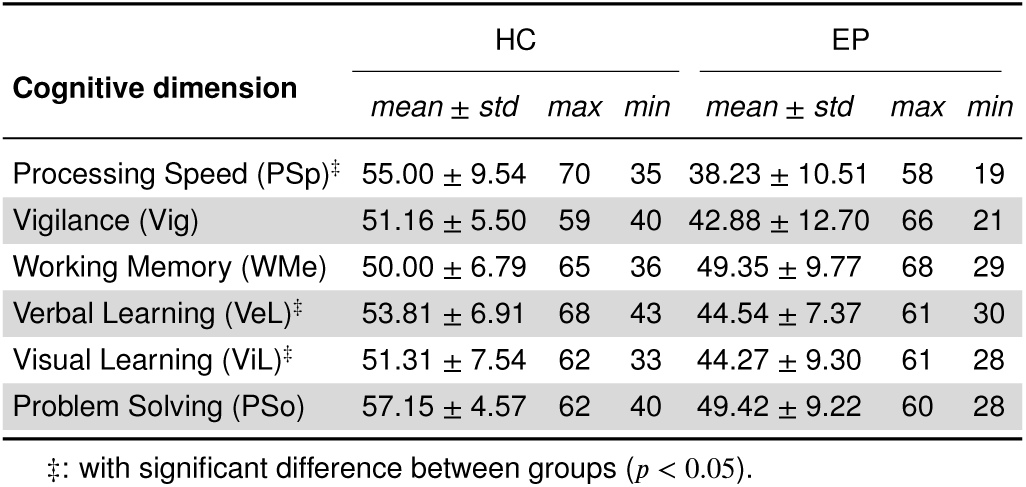
Performance of neurocognition tests.

As pointed out by previous studies [47, 54], the estimation of cognitive level can be done by either classification or regression, that classification task is to manually classify continuous scores into different discrete categories and predict the probability of which category each case should be in, whereas regression is a direct prediction of scores. In this study, for the classification task, we evenly divided the scores between the maximum and minimum values into *n* equal parts, *i*.*e*., *n* categories. It is worth noting that since the maximum and minimum values are different for each cognitive assessment, the interval is also different among the *n* categories. Normally, larger *n* represents a more fine separation of cognitive levels and greater difficulty in prediction.

### 2.3. Proposed Method

#### 2.3.1. 3D-CNN Multi-task Learning Framework

In this study, 3D sMRI arrays were directly used as input for classifications, so we applied 3D-CNN models as a deep learning architecture to encode visual features, similar in previous studies [39, 46, 22]. Instead of dividing the sMRI array into 2D images and using 2D-CNN [45, 24] for feature encoding, 3D-CNN can consider all inputs at once to better capture local features in the 3D space and contribute to the final classification.

To predict both the cognitive level and the probability of EP for each participant, we further introduced a multi-task learning framework. Based on the same visual features extracted by the 3D-CNN, three independent MLP networks were used as individual subbranches for different tasks, including EP classification, cognitive level classification (CLC) and cognitive level regression (CLR). The complete architecture of our 3D-CNN encoder and multi-task learning framework is depicted in Figure 1B and corresponding details are provided in Table 3. The sequential structure of our 3D-CNN encoder was inspired by the previous study on schizophrenia classification [22].

**Table 3.**
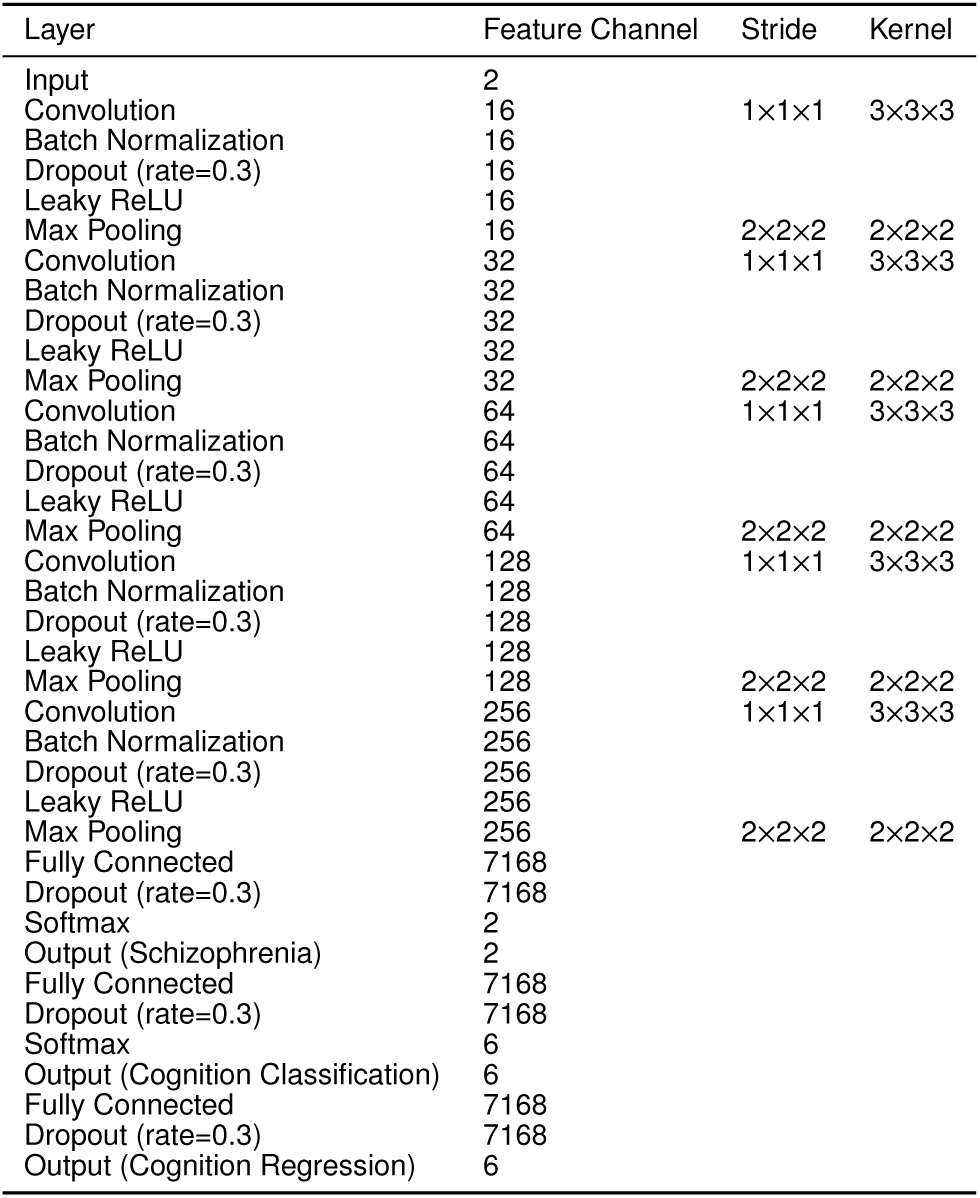
Details of the 3D-CNN architecture

#### 2.3.2. Multi-channel 3D Array Input

We consider the GM and WM probability maps as two different feature channels and make channel concatenations to generate a single 3D array as the input to our model. Unlike previous study [22], where different segmentation components were used as multiple inputs and fed into a model in parallel, our multi-channel 3D array helps to reduce the training parameters and retain all the information from GM and WM. In this case, the dimension of input 3D array will be *H* × *W* × *D* × 2, where *H, W, D* denotes height, width, depth and 2 is the number of channels. The full volume of size 120 × 120 × 120 × 2, rather than smaller volume patches, was used for training and testing. Furthermore, in experiments where only GM or WM is used for training, a single probability map will be replicated once to remain the dimensionality of the input 3D array.

#### 2.3.3. End-to-end Training

Our framework is an end-to-end deep learning system and thus several loss functions were used to train the proposed model for parameter updating. Specifically, for classification tasks (*i*.*e*., EP and cognitive level classification), the conventional cross entropy (CE) loss is used, which is defined as

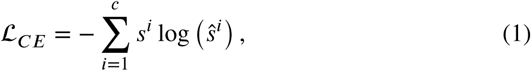

where *s* is the true label, *ŝ* is the prediction, and *c* is the number of class. For the task of cognition regression, the mean square error (MSE) loss is used, which is defined as

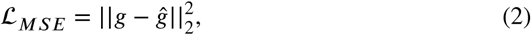

where *g* and *ĝ* denote ground truth label and prediction, respectively. The final loss function is defined as:

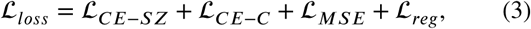

where ℒ_*CE*−*SZ*_ denotes CE loss for EP classification, ℒ_*CE*−*C*_ denotes CE loss for cognitive level classification, ℒ_*MSE*_ denotes MSE loss for cognitive level regression, and ℒ_*reg*_ represents the regularization loss (or weight decay [17]) used to avoid overfitting. As an end-to-end framework, training losses are back-propagated from three multi-task sub-branches to the 3D-CNN, updating the parameters of the entire network with an optimization algorithm (*e*.*g*., Adam [27]). Finally, through minimizing the ℒ_*loss*_, the network could learn a nonlinear mapping from the input 3D sMRI array to EP and cognitive state, enabling EP classification and cognitive estimation for unseen individuals.

## 3. Experiments

### 3.1. Competing Methods

#### 3.1.1. Deep Learning-based Model

Apart from 3D-CNN, we also used a 2D-CNN framework, similar to the model of [24] and [28], for comparison. The latest lightweight 2D convolutional architectures, MNasNet [55], and a cumbersome model, ResNet-18 [21], were used as the feature encoders since they have been commonly used in previous studies [22, 35, 28, 20]. In a 2D-CNN framework, for each participant, image features are extracted slice by slice and concatenated for final classification, which introduces more computational cost than the 3D-CNN model. Furthermore, since 3D-CNNs do not have pre-trained weights like 2D-CNNs, all 3D-CNNs models were trained from scratch. Nevertheless, results are reported for 2D-CNNs with and without pre-trained weights^1^.

#### 3.1.2. Handcrafted Feature-based Machine Learning

To compare with the proposed DL workflow, we also performed the classification tasks with several latest ML methods. The GM and WM probability maps were flattened into feature vectors and the principal component analysis (PCA) was used for dimensionality reduction to produce proper training inputs for ML models. Besides the WM and GM maps, volumetric and surface analysis was also performed with CAT12 toolkit to calculate region of interest (ROI) volumes and cortical surface thickness as handcrafted features for comparison. We adopted the analysis with default settings and obtained 388 ROI volume features and 219 cortical thickness features after filtering out the null values. The Cobra^2^ and neuromorphometircs^3^ were used as ROI atlas. Dimensionality reduction was also performed on handcrafted features to make them the same size as GM/WM-based features. We selected several popular ML models for comparison, including random forest (RF), supported vector machine (SVM) and gradient boost machine (GBM).

### 3.2. Evaluation Metrics and Strategy

We used accuracy, *F*_1_-score, specificity and area under curve (AUC) of receiver operating characteristic (ROC) as the metrics to evaluate the classification performance. Specifically, the *F*_1_-score is the harmonic mean between recall (sensitivity) and precision. The accuracy, *F*_1_-score and specificity are respectively defined as 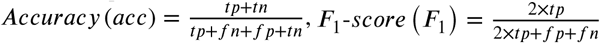 and 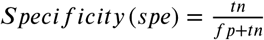, where *tp, fp, tn* and *fn* refer to true positive, false positive, true negative, and false negative, respectively. While *F*_1_-score mainly focus on evaluating prediction performance on positive targets (*i*.*e*., the EP cases), the specificity focus on evaluating the negative ones (*i*.*e*., the healthy cases). All these metrics range from 0 to 1, with higher metrics indicating better predictive performance achieved by the model. In addition, we adopted mean absolute error (MAE) and coefficient of determination (R^2^) as metrics to evaluate regression performance, which is defined as 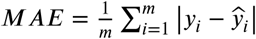 and 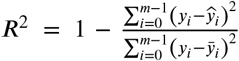, where *m* denotes number of samples, *y* and *ŷ* denote ground truth and prediction, respectively.

Since the size of the data set is relatively small for a deep learning model, we applied a five-fold cross-validation strategy in this study in order to thoroughly evaluate and avoid overfitting. There were 77 3D sMRI arrays after preprocessing. These samples were divided into five parts equally, and one part of them was selected one by one as the test set and the rest as the training set. After that, all metrics are presented as the mean and standard deviation of the five experiments.

### 3.3. Implementation Details

All models were implemented with the Python (version 3.7) programming language and several free Python-based packages. For ML models, the GBM was implemented with a popular lightGBM^4^ framework and other models were implemented using scikit-learn toolkit [41]. The number of estimators in RF model was set as 500 and radial basis function kernel was used in SVM model.

We used PyTorch (version 1.6 stable) as the DL framework to implement all DL-based models. The Adam [27] was used as the optimizer with a starting learning rate of 1e-4, and the learning rate was made to decay by 0.7 after every 60 epochs to help reach optima. Data augmentation (random rotation and flipping) and weight decay of the optimizer (at a rate of 0.02) were used as data set expansion and regularization, respectively, to help prevent overfitting. The batch size was set to 10, and 300 epochs were used. All experiments were conducted on an Ubuntu 18.04 system with two NVIDIA GeForce RTX 2080 Ti graphical processing unit (GPU) and 22 gigabytes memory. The versions of Compute Unified Device Architecture (CUDA) and the driver for the GPU were 10.2 and 460.73.01, respectively. We used a grid search strategy to determine the hyperparameters with learning rates in the range of [1e-3, 1e-4, 1e-5], batch sizes in the range of [4, 8, 10, 12], and weight decay in the range of [0.0, 0.1, 0.2, 0.3, 0.4].

## 4. Results

### 4.1. Cognitive Estimation Performance

In this section, we first evaluate the cognitive estimation performance of the proposed method and competing methods in terms of CLC task. As shown in Table 4, our model achieves better CLC performance in most cases. Specifically, our model obtains the best *F*_1_ score of 70.1% on the two-categorized (*i*.*e*., *n*=2) CLC task. Same results can be observed on the three- and ten-categorized (*n*=3 and *n*=10) CLC that our method outperforms all other counterparts with significant margins. Although in the only case (*n*=5) our method did not get the first place, we still got the second best performance. Based on these results, it can be seen that the proposed DL model was able to classify individuals’ cognitive states into groups using sMRI and achieved promising performance on two-categorized CLC task with higher accuracy than chance.

**Table 4.** Results of *F*_1_-score (%) for cognition classification on average of six dimensions, here *n* denotes number of categories. The best results are in bold.

| Models | $n=2$ | $n=3$ | $n=5$ | $n=10$ |
| --- | --- | --- | --- | --- |
| Volume + SVM | 48.5 ± 11.3 | 31.0 ± 12.5 | 18.8 ± 8.6 | 8.0 ± 4.5 |
| Thickness + SVM | 44.5 ± 9.7 | 29.0 ± 9.4 | 18.1 ± 7.2 | 8.3 ± 4.8 |
| Volume + DNN | 66.2 ± 13.8 | 46.9 ± 10.6 | 31.2 ± 8.7 | 15.6 ± 4.0 |
| Thickness + DNN | 66.6 ± 11.6 | 50.4 ± 7.8 | **33.1 ± 6.9** | 15.0 ± 3.8 |
| Image + RF | 46.4 ± 11.6 | 32.2 ± 10.6 | 17.9 ± 8.7 | 8.9 ± 4.6 |
| Image + SVM | 46.3 ± 11.6 | 30.2 ± 10.0 | 15.2 ± 7.4 | 7.2 ± 4.8 |
| Image + GBM | 47.5 ± 11.5 | 35.0 ± 12.3 | 20.3 ± 9.7 | 8.5 ± 5.1 |
| Image + MNasNet [55]<sup>†</sup> | 66.9 ± 3.4 | 50.6 ± 8.4 | 28.7 ± 8.8 | 15.2 ± 3.7 |
| Image + MNasNet [55]<sup>‡</sup> | 67.8 ± 3.1 | 51.0 ± 7.4 | 29.0 ± 9.5 | 15.6 ± 3.8 |
| Image + ResNet-18 [21]<sup>†</sup> | 63.7 ± 3.2 | 44.0 ± 7.9 | 25.7 ± 6.1 | 14.0 ± 3.0 |
| Image + ResNet-18 [21]<sup>‡</sup> | 67.9 ± 4.9 | 51.1 ± 8.6 | 29.4 ± 7.3 | 15.1 ± 3.8 |
| Image + 3D-CNN (ours) | **70.1 ± 3.5** | **51.9 ± 8.1** | 31.9 ± 7.5 | **16.2 ± 3.7** |

<sup>†</sup>: train from scratch; <sup>‡</sup>: using pre-trained weights.

Furthermore, we demonstrate the classification accuracy for each cognition estimation dimension while *n*=2. As shown in Figure 2, all DL-based models achieved better performance than ML-based models. Although the DL-based models using sMRI images as input performed similarly across the four cognitive dimensions (PSp, Vig, ViL, and PSo), it is noteworthy that our method achieved significant improvements in the WMe and VeL dimensions. Thus, our method performs most convincingly for CLC task in all six dimensions and even achieves an accuracy of more than 80% in some dimensions (PSp, ViL and PSo). Since our method uses sMRI images directly as input without further volumetric and cortical surface based analysis, it achieves both the overall best classification performance and efficiency, both of which are crucial for clinical translations.

**Figure 2:**
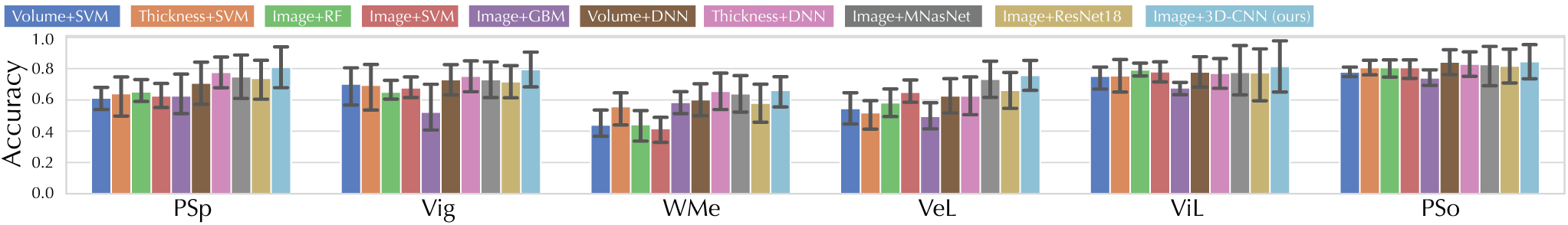
Accuracy of our model in two-categorized CLC task compared with different ML and DL counterparts in six cognitive estimation dimensions. All the DL models shown were trained from scratch.

Besides classification, the regression results for cognitive estimation (*i*.*e*., CLR) are also presented. As shown in Table 5, the CLR performance of all models was worse than expected, even worse than random guesses (R^2^ ≤ 0.0). A possible reason for the poor regression performance may be due to the limited sample size of the data set [9]. By comparing the performance of DL and ML models, it can be seen that DL models generally performed worse than ML models. This suggests that DL models may be more sensitive to the lack of samples [54], and that DL models may be more suitable for classification rather than regression in a sample-limited context.

**Table 5.**
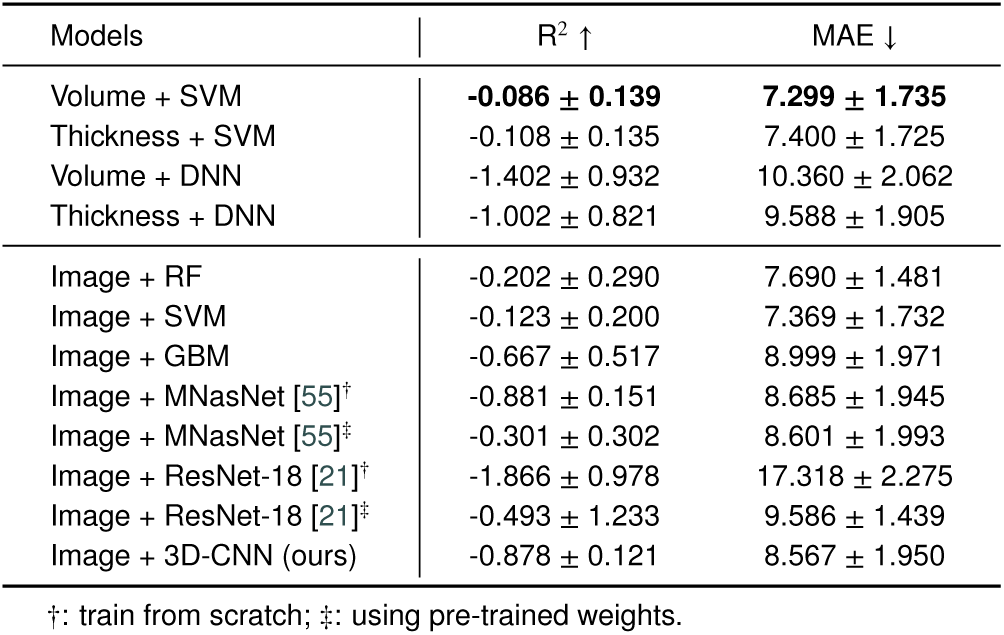
Results for cognition regession on average of six dimensions. The best results are in bold.

| Models | $R^2 \uparrow$ | MAE ↓ |
| --- | --- | --- |
| Volume + SVM | **-0.086 ± 0.139** | **7.299 ± 1.735** |
| Thickness + SVM | -0.108 ± 0.135 | 7.400 ± 1.725 |
| Volume + DNN | -1.402 ± 0.932 | 10.360 ± 2.062 |
| Thickness + DNN | -1.002 ± 0.821 | 9.588 ± 1.905 |
| Image + RF | -0.202 ± 0.290 | 7.690 ± 1.481 |
| Image + SVM | -0.123 ± 0.200 | 7.369 ± 1.732 |
| Image + GBM | -0.667 ± 0.517 | 8.999 ± 1.971 |
| Image + MNasNet [55]<sup>†</sup> | -0.881 ± 0.151 | 8.685 ± 1.945 |
| Image + MNasNet [55]<sup>‡</sup> | -0.301 ± 0.302 | 8.601 ± 1.993 |
| Image + ResNet-18 [21]<sup>†</sup> | -1.866 ± 0.978 | 17.318 ± 2.275 |
| Image + ResNet-18 [21]<sup>‡</sup> | -0.493 ± 1.233 | 9.586 ± 1.439 |
| Image + 3D-CNN (ours) | -0.878 ± 0.121 | 8.567 ± 1.950 |

<sup>†</sup>: train from scratch; <sup>‡</sup>: using pre-trained weights.

### 4.2. Early Psychosis Classification Performance

To evaluate the EP classification performance of our model, we compare it with several latest counterparts [31, 42, 61, 16, 28], of which models were re-implemented based on the settings of the original publications. As shown in Table 6, our proposed model generally outperforms the other five competing methods in all metrics. For instance, our model using solely sMRI images (with GM as input) achieved the best F_1_-score (74.5%) compared to ML-based models using volumetric features (54.1% [31]) and cortical thickness features (55.5% [61] and 60.8% [16]). In addition, our 3D-CNN model also achieves better performance in all metrics compared to 2D-CNN [28], indicating that features are extracted directly from 3D sMRI arrays more efficiently than from 2D slices. Finally, we compared the performance of our model with and without cognitive estimation as a sub-task. By adding cognitive estimation, the accuracy, F_1_ score and specificity were improved by 3.9%, 4.4% and 8.5%, respectively, when GM was used as input. And similar improvements are seen when WM and GM were used as inputs, by 2.9%, 3.7% and 4.8% on the accuracy, F_1_ score and specificity, respectively. Moreover, as shown in Figure 3, the cognitive estimation subtask brought a 4.8% improvement in AUC and also achieved the best classification performance (71.1% on AUC) of all models, further demonstrating its validity. This is consistent with the idea in previous studies that the association between brain abnormalities and cognitive symptoms may exist at a deep and abstract level and thus can be effectively captured by DL methods, leading to enhanced performance in EP classification [43, 58]. These results demonstrate the effectiveness of using 3D-CNN and involving a cognitive estimation subtask for promising EP classification performance.

**Table 6.** Comparison on sMRI-based studies for EP classification. All results are shown in percentage and the best results are highlighted in bold.

| Method | acc | $F_1$ | spe |
| --- | --- | --- | --- |
| Volume features + SVM [31] | 50.8 ± 5.6 | 54.1 ± 8.8 | 53.4 ± 7.3 |
| Volume features + DNN [42] | 65.7 ± 13.4 | 69.0 ± 11.8 | 69.5 ± 13.3 |
| Thickness features + SVM [61] | 58.4 ± 9.0 | 60.8 ± 10.6 | 60.4 ± 10.1 |
| Thickness features + RF [16] | 50.7 ± 13.5 | 55.5 ± 11.7 | 54.6 ± 10.7 |
| sMRI images + 2D-CNN [28] | 68.9 ± 5.6 | 69.4 ± 5.1 | 71.0 ± 5.7 |
| Proposed w/o cognitive estimation<sup>†</sup> | 70.6 ± 4.1 | 70.5 ± 4.1 | 75.3 ± 5.8 |
| Proposed w/ cognitive estimation<sup>†</sup> | 73.5 ± 3.3 | 74.2 ± 3.0 | 80.1 ± 5.1 |
| Proposed w/o cognitive estimation<sup>‡</sup> | 71.0 ± 4.3 | 70.1 ± 4.4 | 73.8 ± 5.9 |
| Proposed w/ cognitive estimation<sup>‡</sup> | **74.9 ± 4.3** | **74.5 ± 4.2** | **82.3 ± 6.3** |

<sup>†</sup>: WM and GM inputs; <sup>‡</sup>: GM input.

**Figure 3:**
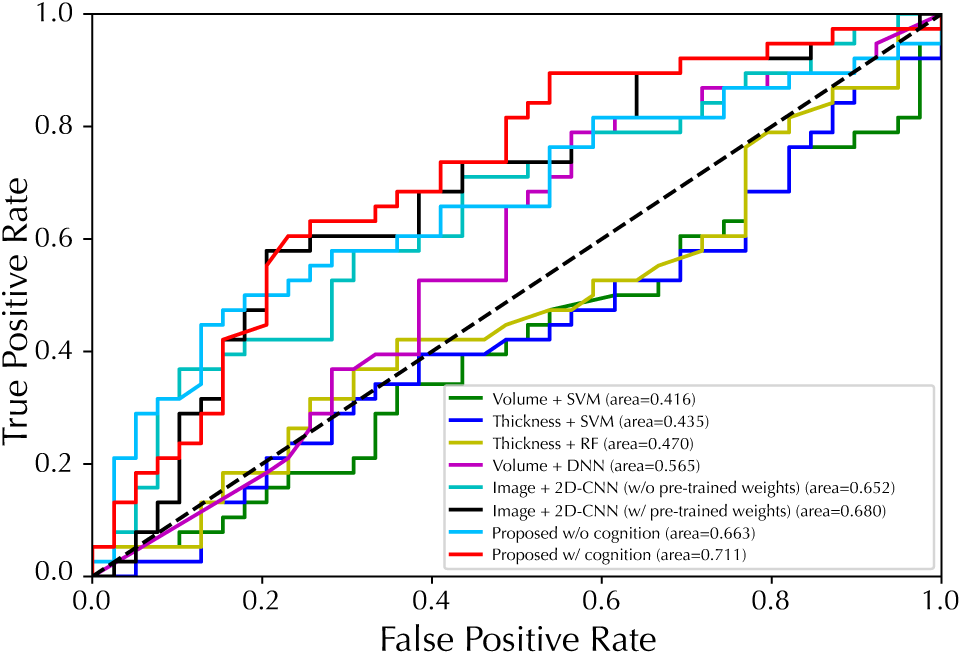
Performance of ROC curves for EP/HC classification with fivefold cross-validation. The proposed model used GM map as input.

## 5. Discussion

In this study, we first evaluate the impact of the CLC subtask on EP classification performance when different numbers of categories are involved. We then study the influence brought by using different sMRI inputs or integrating different cognitive assessment subtasks (CLC or CLR) through several ablation studies. We also present the discriminative regions identified by our model as potential biomarkers for EP classification and CLC, and clarify the potential for clinical translation.

### 5.1. Impact of Cognition Classification Category Quantity

As we hypothesized that the introduction of a cognitive classification task could bring features about individual brain structure to the DL model, it remains unclear whether more classification categories could lead to more discriminative features for EP classification. Therefore, we divided cognitive scores into different number of categories in the CLC subtask and assessed how this would affect classification performance for EP. As shown in Figure 4, the EP classification performance is largely unaffected in terms of F_1_ score and accuracy, while the specificity could be improved when *n* is set to ten. Therefore, in general, introducing a more challenging context in CLC subtask does not bring more discriminative information to the classification of EP. This may be due to the sample limitation in our study, when *n* is set to a large number, some categories may not have a sample at all. However, the improvement in specificity when *n*=10 suggests that a larger number of categories may lead to better EP classification performance in the presence of abundant data.

**Figure 4:**
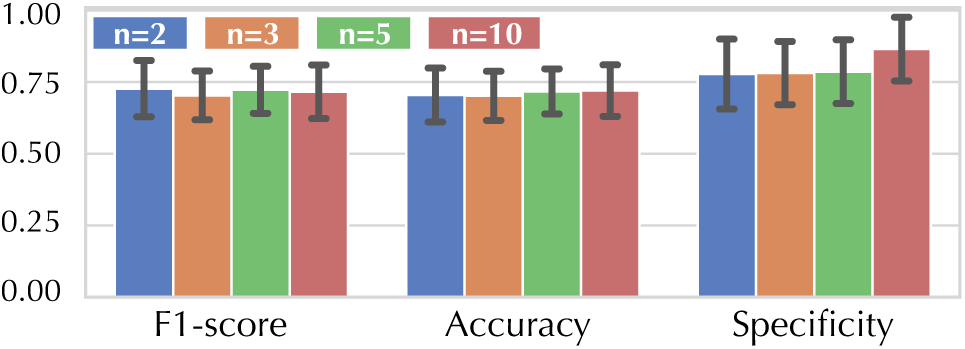
The EP classification performance of our model, when combined with different numbers of categories for CLC. Here *n* denotes number of categories.

### 5.2. Ablation Study

#### 5.2.1. Influence of WM/GM Inputs

WM and GM are two major components of brain tissues, however, it remains unclear how they contribute to the classification of EP in the context of DL. Therefore, we evaluated the effect of using different sMRI images (*i*.*e*., WM or GM images) as input on the EP classification. As shown in Table 7, the model with GM as input outperformed the model with WM as input, with an improved F_1_ score of ≥5.6. This is consistent with previous results that EP causes significant changes in GM [59], while our results further indicate that changes in GM are sufficiently pronounced in EP and can significantly affect the performance of the automated classification tools. Even so, the simultaneous use of WM and GM achieves the best performance in most tasks, confirming the presence of both WM and GM alterations in EP patients. Therefore, despite the best result was obtain when using only GM as input (*i*.*e*., 74.5% for EP + CLR), the inclusion of both GM and WM maps generally resulted in better classification performance for EP.

**Table 7.** Results of *F*_1_ score (%) for EP classification. Here EP denote early psychosis classification. The *n* was set to five for CLC. The best results are in bold.

| Input | Task |  |  |  |
| --- | --- | --- | --- | --- |
|  | EP | EP + CLC | EP + CLR | EP + CLC + CLR |
| WM | 63.2 ± 4.1 | 64.6 ± 4.3 | 64.2 ± 4.4 | 63.5 ± 4.6 |
| GM | 70.1 ± 4.4 | 70.2 ± 5.4 | **74.5 ± 4.2** | 71.2 ± 3.8 |
| WM + GM | **70.5 ± 4.1** | **72.5 ± 4.0** | 73.5 ± 4.0 | **74.2 ± 3.0** |

#### 5.2.2. Influence of Cognition Classification/Regression

We then evaluated how different ways of introducing the cognitive assessment subtask (*i*.*e*., CLC or CLR) contributed to the classification of EP. As shown in Table 8, both CLC and CLR brought improvement on EP classification, while CLR seems to be more effective than CLC. The model incorporating the CLR subtask achieved the best performance on all metrics, with a significant gap compared to the other models. However, performance degrades when CLC is involved in addition to CLR, suggesting that the two subtasks may be incompatible. One possible reason for this is that some discriminative brain regions of the cognitive estimation dimensions may differ from the EP, thus introducing noisy features in the training. In contrast, the regression task did not bring discriminative information, so the features of CLR were more compatible than those of CLC in EP classification. In short, at least for EP classifications, the regression subtask is more informative than classification, but for other diseases the subtask needs to be selected on the merits.

**Table 8.** EP classification performance of our model when introducing different cognitive estimation subtasks, using GM images as input. Here EP denotes early psychosis classification. The *n* was set to five for CLC.

| Model with Task | | | acc | $F_1$ | spe | AUC |
| --- | --- | --- | --- | --- | --- | --- |
| EP | CLC | CLR |  |  |  |  |
| ✓ |  |  | 71.0 ± 4.3 | 70.1 ± 4.4 | 73.8 ± 5.9 | 66.3 ± 3.9 |
| ✓ | ✓ |  | 71.4 ± 3.7 | 70.2 ± 5.4 | 74.4 ± 5.4 | 67.4 ± 4.5 |
| ✓ |  | ✓ | **74.9 ± 4.3** | **74.5 ± 4.2** | **82.3 ± 6.3** | **71.1 ± 4.1** |
| ✓ | ✓ | ✓ | 71.1 ± 3.9 | 71.2 ± 3.8 | 77.0 ± 5.9 | 68.0 ± 4.9 |

### 5.3. Interpretable sMRI Biomarkers and Clinical Potentials

Our proposed framework could potentially identify brain regions that may be associated with psychosis. As shown in Figure 5, we present the attention maps using GradCam++ [8] and GradCam algorithms [50, 15] to illustrate brain structures of importance. Evidently, the entire GM structure contributes mostly to the EP classification, suggesting that more discriminative features are found in GM than in WM, which is in line with the results of better classification performance of the model using GM shown in Table 7. Furthermore, as shown in Figure 5(h), saliency appears in the frontal and temporal lobe regions, as well as putamen, head of caudate nucleus and thalamus. These regions contributed the most to our model in classifying a subject as an EP patient or a healthy subject, suggesting that structural features in these regions are most likely to be discriminative biomarkers for psychosis. Indeed, all of these regions recognized by our model are highly consistent with those reported in previous volumetric and functional studies. Alterations in grey matter, the frontal lobe, putamen, head of caudate nucleus and thalamus were observed in patients with schizophrenia [1, 19, 60, 56] and cognition deficits [2] in group-level volumetric analysis, as well as fMRI studies [30, 3, 14], indicating that the potential of identifying biomarkers from sMRI by DL methods.

**Figure 5:**
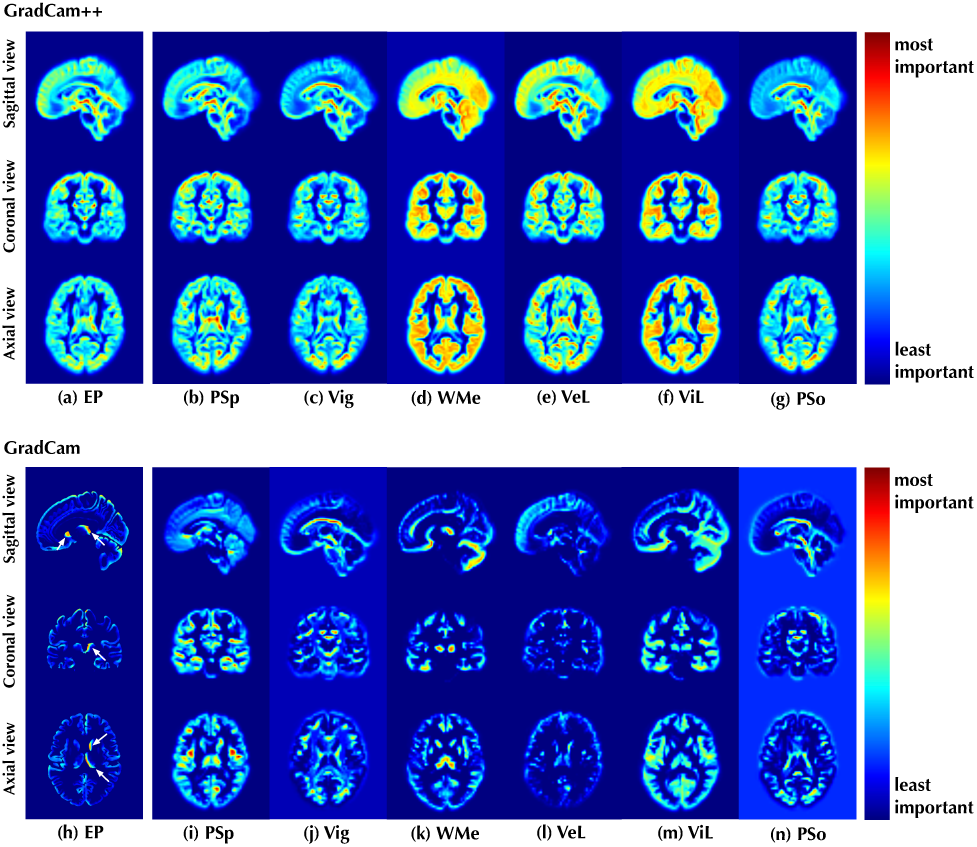
Visualization of the discriminative positions identified by the proposed model on (a, h) EP classification and (b-g, i-n) CLC tasks with attentional weights. The results were shown as the mean of all cases in the data set. We used both WM and GM images as input and *n* was set to two for CLC.

Besides structural biomarkers for psychosis, we also demonstrate the attention maps for CLC in Figure 5(b-g, i-n). Some specific regions are also recognized as discriminative for estimation of cognition level as shown in Figure 5(i-n). Taking working memory as an example, as shown in Figure 5k, the thalamus and cerebellum were highlighted by the DL model with the highest significance, and these regions have also been proved to be associated with working memory function in the previous fMRI studies [5, 18]. Similarly, as shown in Figure 5(m), the highlighted regions of occipital lobe, thalamus and cerebellum for visual learning were also considered associated to visual functions in fMRI studies [44, 7, 40]. Furthermore, as shown in Figure 5(h-n), it is worth noting that the highlighted regions are not only different among the six different cognitive estimation dimensions, but differ significantly from those for EP classification. This could explain why CLC brings less improvement in EP classification than CLR. Since the discriminative regions are different, the model may not be able to coordinate these features to accomplish both tasks simultaneously.

### 5.4. Limitation and Future Work

Although our proposed method achieves improved performance in EP classification and provides biomarkers with a high degree of interpretability, there are still some limitations that may affect the generalizability of our approach. Firstly, the study was conducted at a single site and did not take into account the different ethnic composition and sMRI scanning settings, so multi-site studies are needed for further validation. Secondly, the EP subjects in our study received medication, which may also leads to structural alterations in the brain, thus requiring the use of a non-medicated sample in our future studies to rule out medication interference.

Despite these limitations, our results also lead to many interesting directions for future research. For example, since only EP is studied in this work, whether the cognitive estimation subtask helpful for improving classification performance for other psychiatric disorders could be explored. And, as we demonstrated that implicitly introducing cognitive features in the DL model helps EP classification, the question is raised whether it is better to incorporate such additional features explicitly (*i*.*e*., as input) or implicitly (*e*.*g*., as output) into the workflow. Also, since deep learning and implicit information introduction can enhance classification, with only sMRI as a single input, more other relevant features can be introduced into the model in the same way with the aim of further improving classification performance and providing interpretable evidence to aid clinical translation. Moreover, if validated in larger cohorts of patients at the early phase of psychosis, this approach could open the way to prediction of cognitive deficit in prospective longitudinal study with patients in their prodromal phase.

## 6. Conclusion

In this study, we propose a multitask DL framework for EP classification based on sMRI images. By introducing cognitive estimation as a subtask, the proposed method is able to estimate the cognitive state of an individual and improve the classification performance of EP by an appreciable margin. Experimental results show that our method can not only achieve classification accuracy that exceeds that of the latest similar methods, but also identify discriminative regions in sMRI images as interpretable evidence.

## Data Availability

All data produced in the present study are available upon reasonable request to the authors

## CRediT authorship contribution statement

**Yang Wen**: Conceptualization, Methodology, Software, Investigation, Writing - original draft & review. **Chuan Zhou**: Conceptualization, Methodology, Writing - review & editing, Supervision, Funding acquisition. **Leiting Chen**: Supervision, Funding acquisition. **Yu Deng**: Methodology, Investigation. **Martine Cleusix**: Data curation. **Raoul Jenni**: Data curation, Writing - review & editing. **Philippe Conus**: Data curation. **Kim Q. Do**: Data curation, Writing - review & editing, Resources, Funding acquisition. **Lijing Xin**: Conceptualization, Writing - review & editing, Supervision, Resources, Data curation, Funding acquisition.

## Acknowledgment

This work was supported by Sichuan Science and Technology Program (No.2019YJ0176 / 2019YJ0177 / 2019YFQ-0005), China Scholarship Council, and National Center of Competence in Research (NCCR) “SYNAPSY - The Synaptic Bases of Mental Diseases” from the Swiss National Science Foundation (n° 51AU40_185897 to KQD &PC). We acknowledge access to the facilities and expertise of the CIBM Center for Biomedical Imaging, a Swiss research center of excellence founded and supported by Lausanne University Hospital (CHUV), University of Lausanne (UNIL), EPFL, University of Geneva (UNIGE) and Geneva University Hospitals (HUG).

## Appendix

### A. Participant Recruiting Criteria

Inclusion criteria for EP were a)meeting the psychosis threshold as defined by the Comprehensive Assessment of At-Risk Mental States, b) no antipsychotic medication for >6 months, c) no psychosis related to intoxication or organic brain disease, and d) intelligence quotient >70. Healthy control participants must not a) meet criteria for a DSM Axis 1 and 2 disorder, b) be receiving any current treatment with psychotropic medication, c) have a family history of psychotic spectrum disorder.

### B. Results with Huber Loss

In addition to the MSE loss used for the CLR task, Huber loss [23] was also evaluated, which is defined as

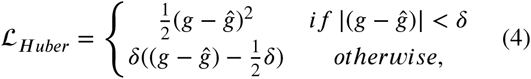

where *g, ĝ* and *δ* denote ground truth label, prediction, and interval, respectively. Compared to the MSE loss, the Huber loss is less sensitive to outliers because it only treats the error as square in the interval of *δ*, which is empirically set to 1.0 in our experiments. As shown in Table 9, Huber loss did not bring significant improvement compared to MSE loss, and in most cases even brought performance degradation. Furthermore, as shown in Table 10, the use of Huber loss in CLR also failed to bring a general improvement to the EP classification task. These results may be due to the fact that the cognitive data we used did not have many outliers to deal with, and a small interval *delta* may lead to gradient loss since some spurious outliers may be incorrectly excluded. Another possible reason is that since there is a new hyperparameter *delta* in the Huber loss, the empirically set value of 1 may not be the optimal value in all cases.

**Table 9.** Results for CLR (in terms of *R*^2^) on average of six dimensions using MSE and Huber loss. The performance gain or loss after using Huber loss compared to MSE loss is represented by ↑ and ↓.

| Models | MSE loss | Huber loss |
| --- | --- | --- |
| Image + MNasNet<sup>†</sup> | -0.881 ± 0.151 | -0.694 ± 0.195 ↑ |
| Image + MNasNet<sup>‡</sup> | -0.301 ± 0.302 | -0.404 ± 0.212 ↓ |
| Image + ResNet-18<sup>†</sup> | -1.866 ± 0.978 | -1.922 ± 1.273 ↓ |
| Image + ResNet-18<sup>‡</sup> | -0.493 ± 1.233 | -0.331 ± 2.019 ↑ |
| Image + 3D-CNN (ours) | -0.878 ± 0.121 | -1.069 ± 0.434 ↓ |

**Table 10.**
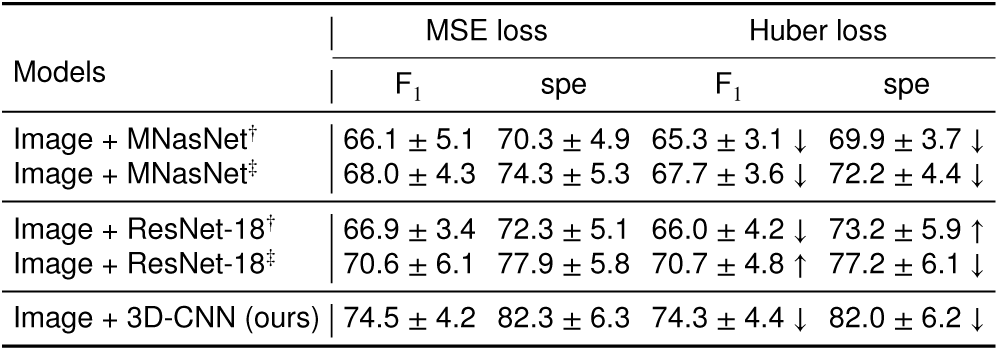
Results for EP classification incorporated with CLR. The performance gain or loss after using Huber loss compared to MSE loss is represented by ↑ and ↓.

**Table 11.** Results and model parameters for CLS (in terms of F_1_) and CLR (in terms of MAE) tasks.

| Models | $F_1 \uparrow$ | MAE ↓ | Parameters |
| --- | --- | --- | --- |
| Image + MNasNet | 67.8 ± 3.1 | 8.6 ± 2.0 | 2.8M |
| Image + ResNet-18 | 67.9 ± 4.9 | 9.6 ± 1.4 | 11.3M |
| Image + 3D-CNN (ours) | 70.1 ± 3.5 | 8.5 ± 2.0 | 1.2M |
| Image + 3D-CNN (1.5× channels) | 70.0 ± 4.9 | 8.8 ± 2.8 | 2.8M |

### C. Computational Costs

We further presented the cognition estimation performance of different models and compared their computational costs. As shown Table 6, the 2D-CNN model has at least two times more parameters compared to the 3D-CNN (ours), but at the same time the performance of cognitive estimation is lower. Instead of extracting features directly from the 3D sMRI volume, the 2D-CNN divided the volume into several slices for feature extraction and combined them into a very large feature embedding for nonlinear projection and final prediction, thus introducing more parameters than the 3D-CNN model. By learning features directly from the 3D volume, the 3D-CNN model not only has fewer parameters than the 2D-CNN model, but also provides better cognitive estimation performance. In addition, we made the parameters of 3D-CNN the same as those of MNasNet by increasing the number of channels of 3D-CNN to 1.5 times of the original one. The modified 3D-CNN model achieved a F_1_ score of 70.0 and MAE of 8.8, which is still better than all 2D-CNN models but worse than the original 3D-CNN model. This performance degradation may be due to an overfitting problem, since we have a relatively small amount of data.

## Footnotes

1 pre-trained weights provided by torchvision package (version 0.7.0)

2 http://cobralab.ca/atlases/

3 https://scalablebrainatlas.incf.org/human/NMM1103

4 https://github.com/microsoft/LightGBM

## References

[1] Alemán-Gómez, Y., Najdenovska, E., Roine, T., Fartaria, M.J., Canales-Rodríguez, E.J., Rovó, Z., Hagmann, P., Conus, P., Do, K.Q., Klauser, P., et al., 2020. Partial-volume modeling reveals reduced gray matter in specific thalamic nuclei early in the time course of psychosis and chronic schizophrenia. Human brain mapping 41, 4041–4061.

[2] Antonova, E., Kumari, V., Morris, R., Halari, R., Anilkumar, A., Mehrotra, R., Sharma, T., 2005. The relationship of structural alterations to cognitive deficits in schizophrenia: a voxel-based morphometry study. Biological psychiatry 58, 457–467.

[3] Argyelan, M., Gallego, J.A., Robinson, D.G., Ikuta, T., Sarpal, D., John, M., Kingsley, P.B., Kane, J., Malhotra, A.K., Szeszko, P.R., 2015. Abnormal resting state fmri activity predicts processing speed deficits in first-episode psychosis. Neuropsychopharmacology 40, 1631–1639.

[4] Baumann, P.S., Crespi, S., Marion-Veyron, R., Solida, A., Thonney, J., Favrod, J., Bonsack, C., Do, K.Q., Conus, P., 2013. T reatment and e arly i ntervention in p sychosis p rogram (tipp-l ausanne): implementation of an early intervention programme for psychosis in s witzerland. Early intervention in psychiatry 7, 322–328.

[5] Bor, J., Brunelin, J., Sappey-Marinier, D., Ibarrola, D., d’Amato, T., Suaud-Chagny, M.F., Saoud, M., 2011. Thalamus abnormalities during working memory in schizophrenia. an fmri study. Schizophrenia research 125, 49–53.

[6] Bora, E., Yalincetin, B., Akdede, B.B., Alptekin, K., 2018. Duration of untreated psychosis and neurocognition in first-episode psychosis: A meta-analysis. Schizophrenia research 193, 3–10.

[7] de Bourbon-Teles, J., Bentley, P., Koshino, S., Shah, K., Dutta, A., Malhotra, P., Egner, T., Husain, M., Soto, D., 2014. Thalamic control of human attention driven by memory and learning. Current biology 24, 993–999.

[8] Chattopadhay, A., Sarkar, A., Howlader, P., Balasubramanian, V.N., 2018. Grad-cam++: Generalized gradient-based visual explanations for deep convolutional networks, in: 2018 IEEE Winter Conference on Applications of Computer Vision (WACV), IEEE. pp. 839–847.

[9] Chauhan, S., Vig, L., De Filippo De Grazia, M., Corbetta, M., Ahmad, S., Zorzi, M., 2019. A comparison of shallow and deep learning methods for predicting cognitive performance of stroke patients from mri lesion images. Frontiers in neuroinformatics 13, 53.

[10] Cortes-Briones, J.A., Tapia-Rivas, N.I., D’Souza, D.C., Estevez, P.A., 2021. Going deep into schizophrenia with artificial intelligence. Schizophrenia Research.

[11] De Marco, M., Beltrachini, L., Biancardi, A., Frangi, A.F., Venneri, A., 2017. Machine-learning support to individual diagnosis of mild cognitive impairment using multimodal mri and cognitive assessments. Alzheimer Disease & Associated Disorders 31, 278–286.

[12] de Filippis, R., Carbone, E.A., Gaetano, R., Bruni, A., Pugliese, V., Segura-Garcia, C., De Fazio, P., 2019. Machine learning techniques in a structural and functional mri diagnostic approach in schizophrenia: a systematic review. Neuropsychiatric disease and treatment 15, 1605.

[13] Fusar-Poli, P., McGuire, P., Borgwardt, S., 2012. Mapping prodromal psychosis: a critical review of neuroimaging studies. European Psychiatry 27, 181–191.

[14] Giraldo-Chica, M., Woodward, N.D., 2017. Review of thalamocortical resting-state fmri studies in schizophrenia. Schizophrenia research 180, 58–63.

[15] Gotkowski, K., Gonzalez, C., Bucher, A., Mukhopadhyay, A., 2020. M3d-cam: A pytorch library to generate 3d data attention maps for medical deep learning. 2007.00453.

[16] Greenstein, D., Weisinger, B., Malley, J.D., Clasen, L., Gogtay, N., 2012. Using multivariate machine learning methods and structural mri to classify childhood onset schizophrenia and healthy controls. Frontiers in psychiatry 3, 53.

[17] Gu, Z., Cheng, J., Fu, H., Zhou, K., Hao, H., Zhao, Y., Zhang, T., Gao, S., Liu, J., 2019. Ce-net: Context encoder network for 2d medical image segmentation. IEEE transactions on medical imaging.

[18] Guell, X., Gabrieli, J.D., Schmahmann, J.D., 2018. Triple representation of language, working memory, social and emotion processing in the cerebellum: convergent evidence from task and seed-based resting-state fmri analyses in a single large cohort. Neuroimage 172, 437–449.

[19] Haijma, S.V., Van Haren, N., Cahn, W., Koolschijn, P.C.M., Hulshoff Pol, H.E., Kahn, R.S., 2013. Brain volumes in schizophrenia: a meta-analysis in over 18 000 subjects. Schizophrenia bulletin 39, 1129–1138.

[20] Hassantabar, S., Zhang, J., Yin, H., Jha, N.K., 2022. Mhdeep: Mental health disorder detection system based on wearable sensors and artificial neural networks. ACM Transactions on Embedded Computing Systems (TECS).

[21] He, K., Zhang, X., Ren, S., Sun, J., 2016. Deep residual learning for image recognition, in: Proceedings of the IEEE conference on computer vision and pattern recognition, pp. 770–778.

[22] Hu, M., Sim, K., Zhou, J.H., Jiang, X., Guan, C., 2020. Brain mri-based 3d convolutional neural networks for classification of schizophrenia and controls, in: 2020 42nd Annual International Conference of the IEEE Engineering in Medicine & Biology Society (EMBC), IEEE. pp. 1742–1745.

[23] Huber, P.J., 1965. A robust version of the probability ratio test. The Annals of Mathematical Statistics, 1753–1758.

[24] Jiang, J., Kang, L., Huang, J., Zhang, T., 2020. Deep learning based mild cognitive impairment diagnosis using structure mr images. Neuroscience letters 730, 134971.

[25] Kern, R.S., Nuechterlein, K.H., Green, M.F., Baade, L.E., Fenton, W.S., Gold, J.M., Keefe, R.S., Mesholam-Gately, R., Mintz, J., Seidman, L.J., et al., 2008. The matrics consensus cognitive battery, part 2: co-norming and standardization. American Journal of Psychiatry 165, 214–220.

[26] Khatri, U., Kwon, G.R., 2020. An efficient combination among smri, csf, cognitive score, and apoe ??4 biomarkers for classification of ad and mci using extreme learning machine. Computational intelligence and neuroscience 2020.

[27] Kingma, D.P., Ba, J., 2015. Adam, a method for stochastic optimization, in: Proceedings of the 3rd International Conference on Learning Representations (ICLR).

[28] Li, Z., Li, W., Wei, Y., Gui, G., Zhang, R., Liu, H., Chen, Y., Jiang, Y., 2021. Deep learning based automatic diagnosis of first-episode psychosis, bipolar disorder and healthy controls. Computerized Medical Imaging and Graphics 89, 101882.

[29] Liu, K., Chen, K., Yao, L., Guo, X., 2017. Prediction of mild cognitive impairment conversion using a combination of independent component analysis and the cox model. Frontiers in human neuroscience 11, 33.

[30] Lord, L.D., Allen, P., Expert, P., Howes, O., Broome, M., Lambiotte, R., Fusar-Poli, P., Valli, I., McGuire, P., Turkheimer, F.E., 2012. Functional brain networks before the onset of psychosis: a prospective fmri study with graph theoretical analysis. NeuroImage: Clinical 1, 91–98.

[31] Lu, X., Yang, Y., Wu, F., Gao, M., Xu, Y., Zhang, Y., Yao, Y., Du, X., Li, C., Wu, L., et al., 2016. Discriminative analysis of schizophrenia using support vector machine and recursive feature elimination on structural mri images. Medicine 95.

[32] Marques, J.P., Kober, T., Krueger, G., van der Zwaag, W., Van de Moortele, P.F., Gruetter, R., 2010. Mp2rage, a self bias-field corrected sequence for improved segmentation and t1-mapping at high field. Neuroimage 49, 1271–1281.

[33] Mohs, R.C., 1999. Cognition in schizophrenia: natural history, assessment, and clinical importance. Neuropsychopharmacology 21, S203–S210.

[34] Nguyen, M., He, T., An, L., Alexander, D.C., Feng, J., Yeo, B.T., Initiative, A.D.N., et al., 2020. Predicting alzheimer’s disease progression using deep recurrent neural networks. NeuroImage 222, 117203.

[35] Noor, M.B.T., Zenia, N.Z., Kaiser, M.S., Al Mamun, S., Mahmud, M., 2020. Application of deep learning in detecting neurological disorders from magnetic resonance images: a survey on the detection of alzheimers disease, parkinsons disease and schizophrenia. Brain informatics 7, 1–21.

[36] Noor, M.B.T., Zenia, N.Z., Kaiser, M.S., Mahmud, M., Al Mamun, S., 2019. Detecting neurodegenerative disease from mri: A brief review on a deep learning perspective, in: International Conference on Brain Informatics, Springer. pp. 115–125.

[37] Nuechterlein, K.H., Green, M.F., Kern, R.S., Baade, L.E., Barch, D.M., Cohen, J.D., Essock, S., Fenton, W.S., Frese III, Ph D F.J., Gold, J.M., et al., 2008. The matrics consensus cognitive battery, part 1: test selection, reliability, and validity. American Journal of Psychiatry 165, 203–213.

[38] Oh, J., Oh, B.L., Lee, K.U., Chae, J.H., Yun, K., 2020. Identifying schizophrenia using structural mri with a deep learning algorithm. Frontiers in psychiatry 11, 16.

[39] Oh, K., Kim, W., Shen, G., Piao, Y., Kang, N.I., Oh, I.S., Chung, Y.C., 2019. Classification of schizophrenia and normal controls using 3d convolutional neural network and outcome visualization. Schizophrenia research 212, 186–195.

[40] Olsson, C.J., Jonsson, B., Nyberg, L., 2008. Learning by doing and learning by thinking: an fmri study of combining motor and mental training. Frontiers in human neuroscience 2, 5.

[41] Pedregosa, F., Varoquaux, G., Gramfort, A., Michel, V., Thirion, B., Grisel, O., Blondel, M., Prettenhofer, P., Weiss, R., Dubourg, V., et al., 2011. Scikit-learn: Machine learning in python. the Journal of machine Learning research 12, 2825–2830.

[42] Pinaya, W.H., Gadelha, A., Doyle, O.M., Noto, C., Zugman, A., Cordeiro, Q., Jackowski, A.P., Bressan, R.A., Sato, J.R., 2016. Using deep belief network modelling to characterize differences in brain morphometry in schizophrenia. Scientific reports 6, 1–9.

[43] Plis, S.M., Hjelm, D.R., Salakhutdinov, R., Allen, E.A., Bockholt, H.J., Long, J.D., Johnson, H.J., Paulsen, J.S., Turner, J.A., Calhoun, V.D., 2014. Deep learning for neuroimaging: a validation study. Frontiers in neuroscience 8, 229.

[44] Poldrack, R.A., Desmond, J.E., Glover, G.H., Gabrieli, J., 1998. The neural basis of visual skill learning: an fmri study of mirror reading. Cerebral Cortex (New York, NY: 1991) 8, 1–10.

[45] Qiu, Y., Lin, Q.H., Kuang, L.D., Zhao, W.D., Gong, X.F., Cong, F., Calhoun, V.D., 2019. Classification of schizophrenia patients and healthy controls using ica of complex-valued fmri data and convolutional neural networks, in: International Symposium on Neural Networks, Springer. pp. 540–547.

[46] Qureshi, M.N.I., Oh, J., Lee, B., 2019. 3d-cnn based discrimination of schizophrenia using resting-state fmri. Artificial intelligence in medicine 98, 10–17.

[47] Rutherford, S., 2020. The promise of machine learning for psychiatry. Biological Psychiatry 88, e53–e55.

[48] Sadeghi, D., Shoeibi, A., Ghassemi, N., Moridian, P., Khadem, A., Alizadehsani, R., Teshnehlab, M., Gorriz, J.M., Nahavandi, S., 2021. An overview on artificial intelligence techniques for diagnosis of schizophrenia based on magnetic resonance imaging modalities: Methods, challenges, and future works. arXiv preprint 2103.03081.

[49] Schwalbe, N., Wahl, B., 2020. Artificial intelligence and the future of global health. The Lancet 395, 1579–1586.

[50] Selvaraju, R.R., Cogswell, M., Das, A., Vedantam, R., Parikh, D., Batra, D., 2017. Grad-cam: Visual explanations from deep networks via gradient-based localization, in: Proceedings of the IEEE international conference on computer vision, pp. 618–626.

[51] Shen, D., Wu, G., Suk, H.I., 2017. Deep learning in medical image analysis. Annual review of biomedical engineering 19, 221–248.

[52] Sommer, I.E., Bearden, C.E., Van Dellen, E., Breetvelt, E.J., Duijff, S.N., Maijer, K., Van Amelsvoort, T., De Haan, L., Gur, R.E., Arango, C., et al., 2016. Early interventions in risk groups for schizophrenia: what are we waiting for? npj Schizophrenia 2, 1– 9.

[53] Spasov, S., Passamonti, L., Duggento, A., Liò, P., Toschi, N., Initiative, A.D.N., et al., 2019. A parameter-efficient deep learning approach to predict conversion from mild cognitive impairment to alzheimer’s disease. Neuroimage 189, 276–287.

[54] Sui, J., Jiang, R., Bustillo, J., Calhoun, V., 2020. Neuroimaging-based individualized prediction of cognition and behavior for mental disorders and health: methods and promises. Biological psychiatry.

[55] Tan, M., Chen, B., Pang, R., Vasudevan, V., Sandler, M., Howard, A., Le, Q.V., 2019. Mnasnet: Platform-aware neural architecture search for mobile, in: Proceedings of the IEEE/CVF Conference on Computer Vision and Pattern Recognition, pp. 2820–2828.

[56] Tao, H., Wong, G.H., Zhang, H., Zhou, Y., Xue, Z., Shan, B., Chen, E.Y., Liu, Z., 2015. Grey matter morphological anomalies in the caudate head in first-episode psychosis patients with delusions of reference. Psychiatry Research: Neuroimaging 233, 57–63.

[57] Vieira, S., Gong, Q.y., Pinaya, W.H., Scarpazza, C., Tognin, S., Crespo-Facorro, B., Tordesillas-Gutierrez, D., Ortiz-García, V., Setien-Suero, E., Scheepers, F.E., et al., 2020. Using machine learning and structural neuroimaging to detect first episode psychosis: reconsidering the evidence. Schizophrenia bulletin 46, 17–26.

[58] Vieira, S., Pinaya, W.H., Mechelli, A., 2017. Using deep learning to investigate the neuroimaging correlates of psychiatric and neurological disorders: Methods and applications. Neuroscience & Biobehavioral Reviews 74, 58–75.

[59] Vita, A., De Peri, L., Deste, G., Sacchetti, E., 2012. Progressive loss of cortical gray matter in schizophrenia: a meta-analysis and metaregression of longitudinal mri studies. Translational psychiatry 2, e190–e190.

[60] Wright, I.C., Rabe-Hesketh, S., Woodruff, P.W., David, A.S., Murray, R.M., Bullmore, E.T., 2000. Meta-analysis of regional brain volumes in schizophrenia. American Journal of Psychiatry 157, 16–25.

[61] Xiao, Y., Yan, Z., Zhao, Y., Tao, B., Sun, H., Li, F., Yao, L., Zhang, W., Chandan, S., Liu, J., et al., 2019. Support vector machine-based classification of first episode drug-naïve schizophrenia patients and healthy controls using structural mri. Schizophrenia Research 214, 11–17.

